# Prevalence of SARS-CoV-2 IgG/IgM antibodies among Danish and Swedish Falck emergency and non-emergency healthcare workers

**DOI:** 10.1101/2020.09.26.20202259

**Authors:** Jannie Laursen, Janne Petersen, Maria Didriksen, Kasper Iversen, Henrik Ullum

## Abstract

**Background:** Knowledge about the COVID-19 outbreak is still sparse especially in a cross-national setting. COVID-19 is caused by a SARS-CoV-2 infection. The aim of the study is to contribute to the surveillance of the pandemic by bringing new knowledge about SARS-CoV-2 seropositivity among healthcare workers and evaluating whether certain job functions is associated with a higher risk of being infected, and to clarify if such association is mediated by the number of individuals that the employees meet during a workday. Finally, we will investigate regional and national differences in seroprevalence.

**Methods:** A bi-national prospective observational cohort study including 3,272 adults employed at Falck in Sweden and Denmark. Participants were tested for SARS-CoV-2 antibodies every second week for a period of 8 weeks from June 22, 2020 until August 10, 2020. Descriptive statistics as well as multivariable logistic regression analyses were applied.

**Results:** Of the 3,272 Falck employees participating in this study, 159 (4.9%) tested positive for SARS-CoV-2 antibodies. The seroprevalence was lower among Danish Falck employees than among those from Sweden (2.8% in Denmark and 8.3% in Sweden). We also found that number of customer or patient contacts during a workday was the most prominent predictor for seropositivity, and that ambulance staff was the most vulnerable staff group.

**Conclusions:** Our study presents geographical variations in seroprevalence within the Falck organization and shows evidence that social interaction is one of the biggest risk factors for getting infected with SARS-CoV-2.

**Key points:** SARS-CoV-2 seroprevalence vary between Denmark and Sweden, between job types and is mostly affected by number of social interactions among Falck healthcare workers

## Introduction

In December 2019, the first case of COVID-19 caused by severe acute respiratory syndrome coronavirus 2 (SARS-CoV-2) was identified. On January 30^th^, 2020, the World Health Organization (WHO) declared the SARS-CoV-2 outbreak to be a public health emergency of international concern[1]. Later it evolved into a pandemic and to date, there have been more than 25 million confirmed cases worldwide, of whom 800,000 have died while infected with the virus[2].

Several strategies for preventing the spread of the virus have been implemented across the world. A strong strategy, which have been widely used, is social distancing. However, for some job functions in the health care sector, this strategy is not feasible to apply. Studies from Denmark and Italy have found a higher seroprevalence among healthcare workers than among the general population[3-5]. Healthcare workers therefore have a particularly high risk of COVID-19.

Falck is a rescue corps employing more than 30,000 healthcare workers worldwide. Most individuals employed by Falck have job functions that put them at risk of being infected with SARS-CoV-2 through interactions with customers or patients. Because such interactions vary between job functions, it is possible that the risk of infection does too. Ambulance staff are presumed to be in high risk of being exposed to individuals infected with SARS-CoV-2, whereas health professionals at clinics mainly interact with the same clients and are therefore not subjected to new people on daily basis. This staff group may therefore have a smaller risk of infection. Moreover, office workers who do not come in contact with either customers or patients may have the lowest risk. Falck also employ part-time firefighters with variable job functions, making it difficult to speculate on the potential level of exposure to SARS-CoV-2 infected individuals. To protect patients, employees and family members of employees from getting infected with SARS-CoV-2, Falck has taken several preventive measures. These varied between Denmark and Sweden as Falck adhered with governmental guidelines. Specific measures taken included hosting telephone or video consultations with patients whenever possible. Moreover, if patients or employees experience any symptoms potentially related to COVID-19, such as fever, coughing, sore throat, headache or sore muscles, they have not been allowed to show up in a clinic. In addition to this, Falck encouraged all patients to have focus on their hygiene and scheduled extra time in between patients for the employees to disinfect. In Denmark, ambulance staff were asked to were masks if they suspected the patient to have COVID-19, while Swedish ambulance staff were asked to were masks at all patient contacts and if they suspected a patient to be infected, they were asked to were complete protective equipment. Falck has 8,000 employees in Denmark and 2,000 in Sweden. For the present study we have tested 2,024 Falck employees in Denmark and 1,248 in Sweden for SARS-CoV-2 antibodies every other week across a period of two months. The aim of the study was to contribute to the surveillance of the pandemic and to bring new knowledge about SARS-CoV-2 seropositivity among healthcare workers by evaluating whether Falck employees with certain job functions have a higher risk of being infected, and to clarify if such association is mediated exclusively by the number of individuals that the employees meet during a workday. Finally, the study investigates regional and national differences in seroprevalence.

### Methods and materials

This is a bi-national prospective observational cohort study including 3,272 individuals above 18 years old. Participants were included on account of being Falck employees. All Falck employees in Denmark (n = 8,000) and Sweden (n = 2,000) were asked to participate in the study and 25.3% of Danish employees agreed, while 62.4% of the Swedish did. Participants were tested for SARS-CoV-2 antibodies every second week for a period of 8 weeks from June 22, 2020 until August 10, 2020.

### Assessing seroprevalence

IgG and IgM antibodies against SARS-CoV-2 were measured in whole blood using the Livzon lateral flow test (Livzon Diagnostics, Zhuhai, Guangdong, China), which we validated with a specificity of 99.54% (95% CI: 98.7-99.9) and a sensitivity of 82.58% (95% CI: 75.7-88.2)[3]. The tests were done by the participants themselves for which they received instructions in writing and on video. The test required that the participants’ put in one drop of blood and two drops of buffer (isotonic saline) in two separate cassettes. After 15 minutes, a conclusive test showed a control line and if IgG or IgM were detected a separate test line for each appeared.

To ensure complete identification of participants with antibodies, test results of participants with only one antibody (IgG or IgM) detected were validated using a different brand of test. For this follow-up test the anti-SARS-CoV-2 (IgG/IgM) POC-test (lateral flow) WONDFO was used. This test was validated in our laboratory at Department of Clinical Immunology, Copenhagen University Hospital, Denmark. It showed a sensitivity of 94.7% (95% CI: 89.8-97.7) and a specificity of 98.7% (95% CI: 97.4-99.4) (unpublished data). The WONDFO test was done by the participants themselves in the same way as the Livzon test.

A positive test was classified as a test indicating the presence of either IgG, IgM or both antibodies.

### Covariates

On the four occasions that participants were tested for SARS-CoV-2 antibodies they also answered a brief questionnaire. In this they reported their job function: ambulance staff, firefighter, healthcare staff, office staff, roadside assistance or field staff. Participants were also asked to report their national region of residence during the study period, and whether they have had an antibody test done outside of this study. Moreover, participants indicated how many individuals they have encountered during workdays on average for the two weeks prior to testing: 0, 1-5, 6-10, 11-20, and more than 20.

### Statistics

Statistical analyses were performed using Enterprise Guide 7.1. Descriptive statistics were conducted to investigate the distribution of study participants, which was presented as frequencies and percentages. National differences in the distribution was examined using *X*^2^-tests for categorial variables. The distribution of participants with a positive test was described using frequencies and percentages. Finally, multivariable logistic regression models were applied to assess the association between job function, number of people contacts and the risk of being tested positive with SARS-CoV-2 antibodies, respectively.

### Ethical statement

Invitations to participate in the study were sent out to all Falck employees in Denmark and Sweden individually through the governmental, personal, password protected email-system *E-boks* in Denmark and *Webropol* in Sweden. To ensure that participants were properly informed before they consented to participate in the study, online live webinars informing about the study were performed for Danish participants. During the webinars participants had the possibility to ask questions directly to the study organizers. It was not a demand from the Scientific Ethics Committee of Sweden that webinars were done, instead Swedish participants received written information and they were given contact information for the study organizer, which they were told to use if they had any questions. The study adheres with the General Data Protection Regulation[6] and the ‘ Scientific Ethical Treatment of Health Science Research Projects’ law[7]. Moreover, participants are pseudonymized and data handling and analysis was conducted by an external statistician with no conflicting interests. Finally, to ensure that participants did not feel pressured into taking part in the study, testing for antibodies was also offered to those employees who did not wish to participate in the study. The study was approved by the Scientific Ethics Committee of Denmark and Sweden, respectively (Denmark: Protocol number: H-20031227 and Sweden: 2020-02862).

## Results

In total, 3,272 (2,024 from Denmark and 1,248 from Sweden) participated in the study. Of these, 64% (n = 2,080) participated in all four rounds of testing, 86% (n = 2,800) participated in at least three rounds and 95% (n=3,096) participated in at least two rounds.

### Characteristics of the study population

There were national differences in the distribution of characteristics between Danish and Swedish participants (Table 1). More men than women participated in Denmark (75.5% men), while the opposite was the case among Swedish participants (32.9% men). In both countries about half of the participants were between the ages 40 and 60 years. A smaller proportion of Danish participants reported more than ten person contacts per day (9.7%) compared to Swedish participants (20.3%). This may be reflected by a smaller proportion of Danish participants being employed as healthcare staff (6.2% in Denmark versus 45.4% in Sweden), while a higher proportion is employed as firefighters (26% in Denmark versus 2.2% in Sweden). Finally, the distribution of employees across the different national regions are in keeping with the size of the regions (Table 1).

**Table 1.**
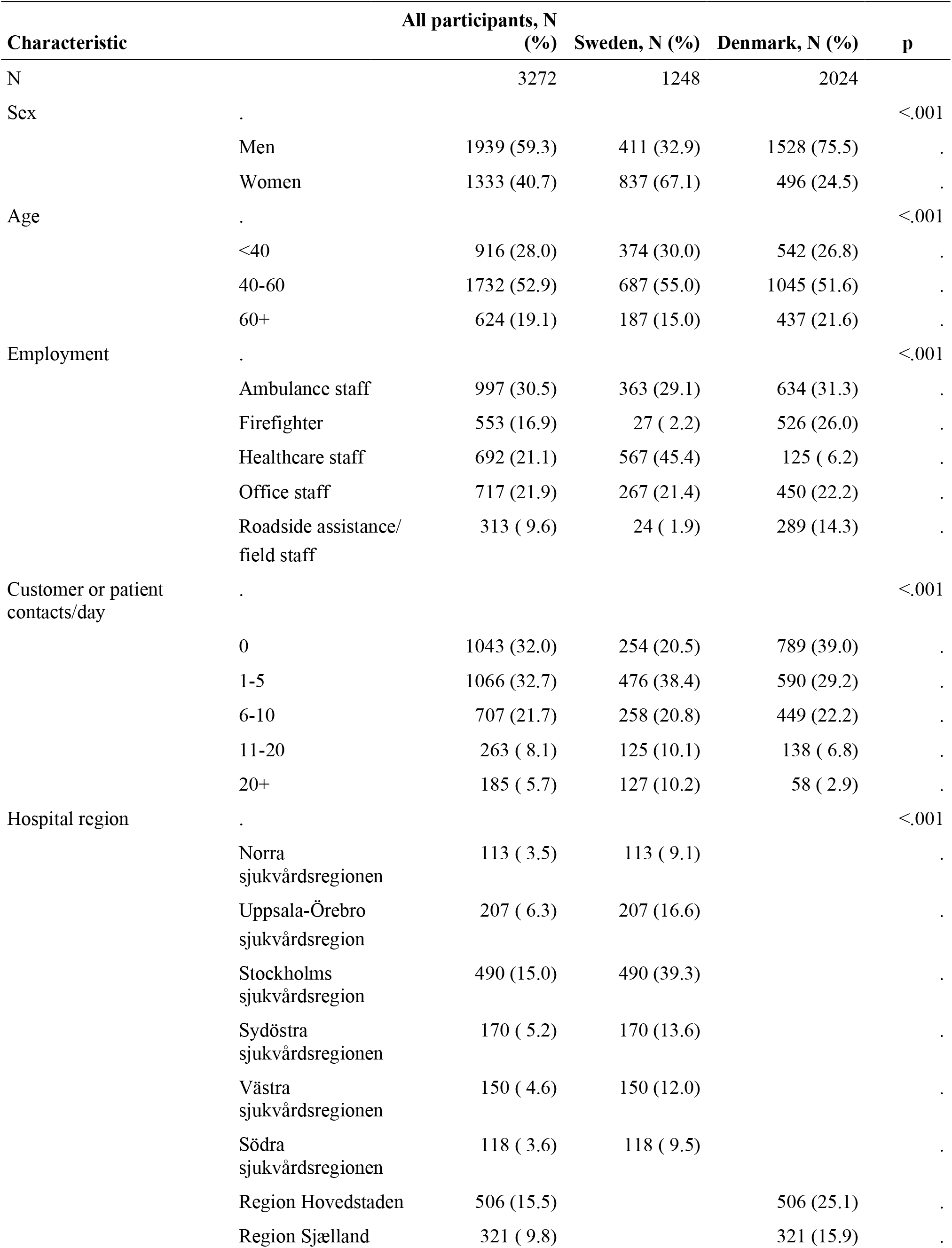
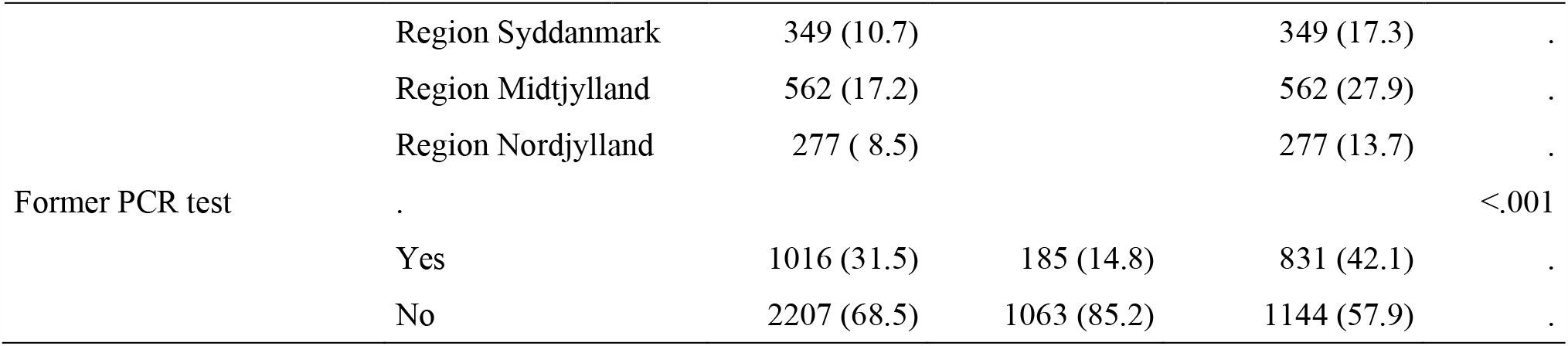
Description of the cohorts in Sweden and Denmark. These descriptions represent in the first test for each employee.

### Proportion of positive immune test

Among the 3,272 included employees, 29 (0.9%) did not have a valid test result. Because of the lower sensitivity compared to specificity a person was considered positive if they had a least one positive test: After the first test 3.3% (n=107) was tested positive, after second test 4,1% (n=133) was tested positive, after three test 4,7% (n=153) and after fourth test 159 (4.9%) were tested positive for SARS-CoV-2 antibodies, corresponding to 2.8% of Danish participants and 8.3% of Swedish participants (Table 2). The group of participants aged 60 years and above had the lowest proportion of seropositivity. Ambulance staff had the highest proportion in both countries, whereas firefighters had the lowest. Among Swedish participants it seems that the proportion of seropositivity increases with the number of people contacts during a workday. Similar is not observed among Danish participants (Table 2).

**Table 2.** Number and proportion of employees with at least one positive test and its dependence of employee characteristics

| Characteristic |  | Number with a positive test result (%) | Sweden Positive N (%) | Denmark Positive N (%) |
| --- | --- | --- | --- | --- |
| Sex | Men | 77 ( 4.0) | 34 ( 8.4) | 43 ( 2.8) |
|  | Women | 82 ( 6.2) | 69 ( 8.3) | 13 ( 2.6) |
| Age | <40 | 49 ( 5.4) | 33 ( 9.0) | 16 ( 3.0) |
|  | 40-60 | 90 ( 5.2) | 60 ( 8.8) | 30 ( 2.9) |
|  | 60+ | 20 ( 3.2) | 10 ( 5.4) | 10 ( 2.3) |
| Employment | Ambulance staff | 79 ( 8.0) | 53 (14.7) | 26 ( 4.1) |
|  | Firefighter | 9 ( 1.6) | 1 ( 3.8) | 8 ( 1.5) |
|  | Healthcare staff | 33 ( 4.8) | 31 ( 5.5) | < 3 |
|  | Office staff | 27 ( 3.8) | 16 ( 6.1) | 11 ( 2.5) |
|  | Roadside assistance/field staff | 11 ( 3.5) | < 3 | 9 ( 3.1) |
| Customer or patient contacts/day | 0 | 27 ( 2.6) | 17 ( 6.7) | 10 ( 1.3) |
|  | 1-5 | 50 ( 4.7) | 28 ( 5.9) | 22 ( 3.8) |
|  | 6-10 | 40 ( 5.7) | 22 ( 8.5) | 18 ( 4.0) |
|  | 11-20 | 20 ( 7.6) | 16 (12.9) | 4 ( 2.9) |
|  | 20+ | 21 (11.3) | 19 (14.8) | < 3 |
| Hospital region | Norra sjukvårdsregionen |  | 0 |  |
|  | Uppsala-Örebro sjukvårdsregion |  | 11 ( 5.4) |  |
|  | Stockholms sjukvårdsregion |  | 62 (12.8) |  |
|  | Sydöstra sjukvårdsregionen |  | 16 ( 9.5) |  |
|  | Västra sjukvårdsregionen |  | 11 ( 7.4) |  |
|  | Södra sjukvårdsregionen |  | 3 ( 2.6) |  |
|  | Region Hovedstaden |  |  | 12 ( 2.4) |
|  | Region Sjælland |  |  | 12 ( 3.8) |
|  | Region Syddanmark |  |  | 9 ( 2.6) |
|  | Region Midtjylland |  |  | 18 ( 3.2) |
|  | Region Nordjylland |  |  | 5 ( 1.8) |

The multiple logistic regression analyses identified a dose-response trend between average number of people contacts during a workday and risk of being tested positive for SARS-CoV-2 antibodies. The estimates do not change when adjusted for age and sex, however when also region is included as a covariate, the ORs attenuate, but remain statistically significant. In the full model including information on sex, age, region and type of employment, it appears that the risk of seropositivity is doubled in employees with 11-20 contacts a day compared to zero contacts per day (OR = 2.3, 95% CI: 1.2-4.6), while the risk is tripled in employees with more than 20 contacts a day (OR = 2.9, 95% CI: 1.5-5.8) (Table 3).

**Table 3:** Logistic regression of risk of positive immune covid-19 test

|  | Unadjusted |  | Adjusted for<br>age and sex |  | Adjusted for<br>age, sex and region |  | Full model* |  |
| --- | --- | --- | --- | --- | --- | --- | --- | --- |
|  | OR (95% CI) | P | OR (95% CI) | P | OR (95% CI) | P | OR (95% CI) | P |
| **Number of customer or**<br>**patient contacts a day** |  | 0.00 |  | 0.00 |  | 0.00 |  | 0.02 |
| 0 (*reference*) | 1 |  | 1 |  | 1 |  | 1 |  |
| 1-5 | 1.8 (1.1 - 3.0) |  | 1.9 (1.2-3.0) |  | 1.6 (1.0 - 2.7) |  | 1.4 (0.8-2.5) |  |
| 6-10 | 2.2 (1.4 - 3.7) |  | 2.3 (1.4-3.9) |  | 2.1 (1.3 - 3.5) |  | 1.7 (0.9-3.0) |  |
| 11-20 | 3.1 (1.7 - 5.6) |  | 3.3 (1.8-5.9) |  | 2.6 (1.4 - 4.8) |  | 2.3 (1.2-4.6) |  |
| 20+ | 4.8 (2.6 - 8.6) |  | 4.6 (2.6-8.4) |  | 3.4 (1.8 - 6.4) |  | 2.9 (1.5-5.8) |  |
| **Employment type** |  | 0.00 |  | 0.00 |  | 0.00 |  | 0.06 |
| Office staff / field staff<br>(*reference*) | 1 |  | 1 |  | 1 |  | 1 |  |
| Ambulance staff | 2.2 (1.4 - 3.4) |  | 2.7 (1.7 - 4.3) |  | 2.1 (1.3 - 3.4) |  | 1.4 (0.8 - 2.6) |  |
| Firefighter | 0.4 (0.2 - 0.9) |  | 0.6 (0.3 - 1.3) |  | 0.7 (0.3 - 1.6) |  | 0.6 (0.2 - 1.3) |  |
| Healthcare staff | 1.3 (0.8 - 2.1) |  | 1.2 (0.7 - 2.1) |  | 1.0 (0.6 - 1.8) |  | 0.8 (0.4 - 1.5) |  |
| Roadside assistance | 0.9 (0.5 - 1.9) |  | 1.3 (0.6 - 2.8) |  | 1.5 (0.7 - 3.2) |  | 1.0 (0.5 - 2.3) |  |

\*Model includes sex, age, national region, number of customer/patients contacts a day, and employment type

Compared to office staff/field staff, the crude model showed that ambulance staff had the highest risk of getting a positive test response (OR = 2.2, 95% CI: 1.4-3.4). Healthcare staff also had an increased risk compared to office staff/field staff (OR = 1.3, 95% CI: 0.8-2.1), while firefighters and roadside assistance had a lower risk of infection (OR = 0.4, 95% CI: 0.2-0.9; OR = 0.9, 95% CI: 0.5-1.9, respectively). When adjusted for age, sex and region and the OR’ s attenuated. Applying the full model, which also included number of people contacts a day, ORs attenuated further and became statistically insignificant (Table 3). Because of the low seroprevalence among Danish participants, follow-up analyses only including Swedish participants were done. These showed similar results (Table 4).

**Table 4:** Logistic regression of risk of positive immune covied-19 test

|  | <u>Only including Swedish participants</u> |  | Adjusted for<br>age and sex |  | Adjusted for<br>age, sex and region in<br>Sweden |  | Full model* |  |
| --- | --- | --- | --- | --- | --- | --- | --- | --- |
|  | Unadjusted |  |  |  |  |  |  |  |
|  | OR (95% CI) | P | OR (95% CI) | P | OR (95% CI) | P | OR (95% CI) | P |
| **Number of customer or**<br>**patient contacts a day** |  | 0.00 |  | 0.00 |  | 0.00 |  | 0.03 |
| 0 (*reference*) | 1 |  | 1 |  | 1 |  | 1 |  |
| 1-5 | 0.9 (0.5 - 1.6) |  | 0.9 (0.5 - 1.7) |  | 1.0 (0.5 - 1.9) |  | 0.9 (0.4 - 1.8) |  |
| 6-10 | 1.3 (0.7 - 2.5) |  | 1.3 (0.7 - 2.5) |  | 1.4 (0.7 - 2.8) |  | 1.2 (0.5 - 2.5) |  |
| 11-20 | 2.1 (1.0 - 4.2) |  | 2.1 (1.0 - 4.3) |  | 2.3 (1.1 - 4.7) |  | 2.0 (0.9 - 4.5) |  |
| 20+ | 2.4 (1.2 - 4.8) |  | 2.4 (1.2 - 4.8) |  | 2.8 (1.4 - 5.7) |  | 2.3 (1.0 - 5.1) |  |
| **Employment type** |  | 0.00 |  | 0.00 |  | 0.06 |  | 0.23 |
| Office staff / field staff<br>(*reference*) | 1 |  | 1 |  | 1 |  | 1 |  |
| Ambulance staff | 2.6 (1.5 - 4.7) |  | 2.9 (1.6 - 5.2) |  | 2.3 (1.2 - 4.3) |  | 1.8 (0.9 - 3.7) |  |
| Firefighter | 0.6 (0.1 - 4.9) |  | 0.7 (0.1 - 6.0) |  | 0.7 (0.1 - 6.1) |  | 0.4 (0.0 - 3.8) |  |
| Healthcare staff | 0.9 (0.5 - 1.7) |  | 0.9 (0.5 - 1.7) |  | 1.2 (0.6 - 2.2) |  | 1.0 (0.5 - 2.1) |  |
| Roadside assistance | 1.4 (0.3 - 6.5) |  | 1.7 (0.4 - 8.2) |  | 1.2 (0.3 - 6.1) |  | 1.1 (0.2 - 5.4) |  |

\*Model includes sex, age, national region, number of customer/patients contacts a day, and employment type

## Discussion

Of the 3,272 Falck employees participating in this study, 159 (4.9%) tested positive for SARS-CoV-2 antibodies. The seroprevalence was lower among Danish Falck employees than among those from Sweden (2.8% in Denmark and 8.3% in Sweden). We also found that number of customer or patient interactions during a workday was the most prominent predictor for seropositivity.

It is plausible that the national variance in seroprevalence between the two countries is a result of different governmental strategies for dealing with the pandemic. The seroprevalences of 2.8% and 8.3% observed among Danish and Swedish Falck employees, respectively are higher than those observed among Danish (1.7%)[8] and Swedish (6.8%)[9] otherwise healthy blood donors. Blood donors represent an age and sex distribution similar to that of the background population between the ages 18 and 65. An explanation for part of the increase in the seroprevalence is that this expected to increase with time.

Before the study commenced, we hypothesized that employees could be divided into risk groups dependent on the suspected number of people contacts during a workday, with a higher number indicating a higher risk. The present results validate this, as analyses revealed a dose-response trend in the association between number of customer or patient interactions and risk of seropositivity. Furthermore, analyses showed that customer or patient interactions had a higher impact on the risk than job function did. We also found that ambulance staff had the highest risk of seropositivity. The OR attenuated when adjusted for sex, age, and national region. The OR attenuated even further and became statistically insignificant when number of people contacts a day was added as a covariate in the statistical model. Therefore, it is plausible that the observed increased risk is explained by a related high level of customer or patient interaction, but it may also be that work exposure for ambulance staff confer a particular risk as ambulance staff cannot reject patients with possible symptoms of COVID-19. Even though Danish ambulance staff were only asked to wear masks when they came in contact with a potential COVID-19 sufferer, while Swedish ambulance staff were asked to wear masks at all patient contacts we found a significantly higher seroprevalence among Swedish ambulance staff (14.7%) than among Danish (4.1%). This may be evidence that the masks carried by staff did not protect against infection. A way to potentially increase the efficiency is to ask patients to wear masks as well.

In support of our finding, a recently published study including healthcare workers from the Capital Region of Denmark found that paramedics had highest seroprevalence out of all hospital staff (4.95% of paramedics tested positive versus 4.04% of all hospital staff)[3]. In the same study it was also found that the seroprevalence was significantly higher in men compared to women[3], which the present study do not support. This may be explained by the fact that there are more women working as ambulance staff in Sweden, while there are more men in Denmark. Moreover, the present study shows that a smaller proportion of employees above 60 years old test positive for antibodies. This may be explained by the elderly being aware of their increased risk of more severe COVID-19 illness and therefore taking more personal precautions to avoid infection.

A specific strength of the present study is the inclusion of participants from different countries and from different regions within these. It is also a strength that the participants were tested for antibodies several times across the study period, which ensured a more accurate estimation of the seroprevalence. This is important as antibodies develop up to 19 days after having COVID-19[10]. Another methodological strength is the fact that participants were not selected due to experience of COVID-19 symptoms.

Limitations of the study is the self-reported nature of data. Participants had to perform the antibody tests themselves and report the results back to the study organizers. This may have caused misclassification of cases. However, if such bias exists it is likely to be random and therefore not affect study results. Moreover, the relatively low sensitivity of the test (82.58%) has potentially caused an underestimation of the seroprevalence. Testing participants multiple times may have reduced the level of underestimation. The potential underestimation was further reduced by additional testing of participants who tested positive for only one of the two SARS-CoV-2 antibodies IgM or IgG. Another limitation to the study is the number of participants. Since the seroprevalence is low, the statistically insignificant findings may be an artefact of reduced statistical power. Moreover, because of the low seroprevalence and because the infection rate has been low during the study period, we did not have statistical power to investigate the development of the seroprevalence across time or to test for interactions. Descriptive statistics presenting the distribution of seropositivity among study participants, seem to present a trend among Swedish employees showing that the more people contact an employee encounter during a workday, the higher the proportion of employees with a positive test. We do not observe a similar trend among Danish participants. However, this is likely to be explained by the low number of Danish employees with more than 10 contacts a day.

To conclude, this is the first bi-national investigation of SARS-CoV-2 seroprevalence among healthcare workers. The Falck concern employs many people with different job functions and therefore represent groups in different risks of getting infected with SARS-CoV-2. Findings from this study represent an important contribution to surveillance of seropositivity in society and to the understanding of how this virus spreads. Such knowledge is imperative in constructing the most appropriate public health policies for dealing with the pandemic. Our study clearly shows that social interaction with customers or patients is the biggest risk factor for getting infected with SARS-CoV-2 in our study population. Moreover, we observed a higher seroprevalence among Falck employees than among the background population in both countries, and we found a significant variance in seroprevalence between employees in Denmark and Sweden.

## Data Availability

Data can be made available upon request to Professor Henrik Ullum

## Funding

This work was supported by the Lundbeck Foundation [grant number R349-2020-1172].

## Acknowledgements

We wish to thank Falck employees Regine Mobech, Mette Wejs Bojsen and Jakob Riis for their important contribution to this study. Further, we want to express our appreciation to the Falck employees who participated in the study.

## Notes

### Competing Interest Statement

The authors have declared no competing interest.

### Author Declarations

The study was approved by the Scientific Ethics Committee of Denmark and Sweden, respectively (Denmark: Protocol number: H-20031227 and Sweden: 2020-02862).

